# Rare variants in SLC34A3 explain missing heritability of urinary stone disease

**DOI:** 10.1101/2022.12.02.22283024

**Authors:** Omid Sadeghi-Alavijeh, Melanie MY Chan, Shabbir Moochhala, Genomics England Research Consortium, Sarah Howles, Daniel P. Gale, Detlef Böckenhauer

**Author notes:** Corresponding author: Detlef Böckenhauer. DPG and DB contributed equally.

## Abstract

Urinary stone disease (USD) is a major health burden affecting >10% of the UK population at some time. While stone disease is strongly associated with lifestyle, genetic factors also predispose to USD: common genetic variants at multiple loci from genome-wide association studies account for 5% of the estimated 45% heritability of the disorder. We investigated the extent to which rare genetic variation contributes to the unexplained heritability of USD.

Among participants of the UK 100,000 genome project, we identified 374 unrelated individuals assigned diagnostic codes indicative of USD. We performed whole genome gene-based variant burden testing and polygenic risk scoring against a control population of 24,930 genetic ancestry matched controls.

We observed (and replicated in an independent dataset) exome-wide significant enrichment (P=2.61×10-07) of monoallelic rare, predicted damaging variants in SLC34A3 (previously associated with autosomal recessive hereditary hypophosphataemic rickets with hypercalciuria) present in 19 (5%) cases compared with 1.6% of controls. The risk of USD with a monoallelic SLC34A3 variant (OR=3.75, 95% CI 2.27-5.91) was greater than the top decile of polygenic risk (OR=2.31, 95% CI 1.12-3.51). Addition of the SLC34A3 variant binary to a linear model including polygenic score increased the estimated variance explained, increasing the liability adjusted pseudo-R2 from 5.1% to 14.2%. We also observed significant association at OR9K2, an olfactory receptor, but this signal was not replicated.

In this cohort rare variants in SLC34A3 were the most important genetic risk factor for USD, with levels of pathogenicity intermediate between the fully penetrant rare variants linked with Mendelian disorders and the weaker effects of common variants associated with USD. These findings explain some of the heritability unexplained by prior common variant GWAS.

## Introduction

Urinary stone disease (USD) is a significant clinical and societal health burden affecting roughly 10% of the population at some point in their life^1^. The prevalence is increasing and there are now over 80,000 hospital episodes per year in the UK^2^. Consequently, the health economic burden is substantial, estimated around £250,000,000 in England per year for the initial stone treatment alone^2^. In the USA, the annual cost for USD in 2000 was calculated as almost $3 billion and estimated to reach $4 billion by 2030^3^. Moreover, there is a strong association between kidney stones and the development of chronic kidney disease (CKD), further adding to the burden from USD ^4,5^.

The etiology of USD is multifactorial, with genetic and environmental factors implicated. There is a strong association between affluence of a society and the prevalence of USD, likely reflecting Western lifestyle habits that include a high salt and animal protein intake^6^. Yet, there is also a strong genetic contribution: a family history is seen in up to 65% of patients with USD with the heritability of stone disease estimated to be as high as 45%^7–10^. Indeed, a strong family history of kidney stone disease can confer a >50 times risk in an individual^11,12^. At a polygenic level, multiple genome wide association studies (GWAS) have been conducted in multi-ancestry populations with greater than fifteen independent loci reported, accounting for roughly 5% heritability^13^. Moreover, there is increasing realization that the burden of monogenic causes of USD is considerable: in two recent studies up to 20% of subjects with USD were considered to have a monogenic cause for their disease^14,15^.

Identification of underlying genetic factors is important, as it facilitates targeted treatment and specific prognostic and genetic counselling^16^, but the gap between the contribution of the known polygenic risk factors and the observed heritability suggests that important genetic contributors to USD remain to be identified.

The 100,000 Genome Project (100KGP) is a pilot project to assess the utility of whole genome sequencing (WGS) in rare disease diagnosis in routine healthcare^17^. This project’s research arm provides an opportunity to correlate genomic information from participants with their clinical phenotype, as documented in Human Phenotype Ontology (HPO) codes. We therefore aimed to investigate the contribution to nephrolithiasis of rare genetic variants (which are not ascertained by previous GWAS) by performing whole genome gene-based variant burden studies in participants with HPO codes for nephrocalcinosis and/or USD to identify and quantify genetic contributors to the missing heritability of stone disease.

## Methods

We utilized the Genomics England dataset (v15)^18^, which contains WGS data, details of clinical phenotypes encoded using Human Phenotype Ontology (HPO) terms^19^ and National Health Service (NHS) hospital records, collected for more than 90,259 cancer and rare disease patients (see Data availability) as well as their unaffected relatives to generate the cohorts. Ethical approval for the 100KGP was granted by the Research Ethics Committee for East of England – Cambridge South (REC Ref14/EE/1112). Figure S1 details the study workflow. We searched for participants recruited with a primary diagnosis of USD or whose clinical information included HPO and/or Hospital Episode Statistics (HES) codes related to USD (a full list of searched codes is provided in table S1)

All cases recruited for USD had been assessed in the clinical interpretation arm of the 100KGP. This involved ascertainment of variants in an expert curated panel of 29 USD genes with multi-disciplinary review and application of American College of Molecular Genetics (ACMG) criteria to determine pathogenicity^20^. The control cohort consisted of 27,660 unaffected relatives of non-renal rare disease participants, excluding those with HPO terms and/or hospital episode statistics (HES) data consistent with USD or secondary causes of USD, kidney disease or kidney failure. By utilizing a case-control cohort sequenced on the same platform, we aimed to minimize confounding by technical artefacts.

Whole genome sequencing was performed by Genomics England, as described previously^17^.

### gVCF annotation and variant-level quality control

gVCFs were aggregated using gvcfgenotyper (Illumina, version: 2019.02.26) with variants normalized and multi-allelic variants decomposed using vt^21^ (version 0.57721). Variants were retained if they passed the following filters: missingness ≤ 5%, median depth ≥ 10, median GQ ≥ 15, percentage of heterozygous calls not showing significant allele imbalance for reads supporting the reference and alternate alleles (ABratio) ≥ 25%, percentage of complete sites (completeGTRatio) ≥ 50% and P value for deviations from Hardy-Weinberg equilibrium (HWE) in unrelated samples of inferred European ancestry ≥ 1×10-5. Male and female subsets were analyzed separately for sex chromosome quality control. Per-variant minor allele count (MAC) was calculated across the case-control cohort, MAC is defined as the number of minor alleles counted for each marker. Annotation was performed using Variant Effect Predictor^22^ (VEP, version 98.2) including CADD^23^ (version 1.5), and allele frequencies from publicly available databases including gnomAD^24^ (version 3) and TOPMed^25^ (Freeze 5). Variants were filtered using bcftools^26^ (version 1.11).

### Cohort rationale

Given the small number of recruited cases, we chose to jointly analyze individuals from diverse ancestral backgrounds, thereby preserving sample size and boosting power. To mitigate confounding due to population structure whilst using this mixed ancestry approach we employed two strategies, as previously described^27^. First, we carried out ancestry-matching of cases and controls using weighted principal components and second, utilized a generalized logistic mixed model to account for relatedness between individuals.

### Relatedness estimation and principal components analysis

A set of 127,747 high quality autosomal LD-pruned biallelic single nucleotide variants (SNVs) with a minor allele frequency (MAF) > 1% was generated using PLINK^28^ (v1.9), MAF is defined as the frequency at which the second most common allele occurs in a given population. SNVs were included if they met all the following criteria: missingness < 1%, median GQ ≥ 30, median depth ≥ 30, AB Ratio ≥ 0.9, completeness ≥ 0.9. Ambiguous SNVs (AC or GT) and those in a region of long-range high LD (Linkage Disequilibrium) were excluded. LD pruning was carried out using an xsr^2^ threshold of 0.1 and window of 500kb. SNVs out of HWE in any of the AFR, EAS, EUR or SAS 1000 Genomes populations were removed (pHWE < 1 ×10-5). Using this variant set, a pairwise kinship matrix was generated using the PLINK2 implementation of the KING-Robust algorithm^29^ and a subset of unrelated samples was ascertained using a kinship coefficient threshold of 0.0884 (2nd degree relationships). Ten principal components were generated using PLINK2 for ancestry-matching and as covariates in the association analyses.

### Ancestry-matching of cases and controls

Given the mixed-ancestry composition of the cohort we employed a case-control ancestry-matching algorithm to optimize genomic similarity and minimize the effects of population structure as previously described^27^ with each case having to match a minimum of two controls to be included in the final cohort. 374 cases and 24930 controls remained for analysis (Fig. S2).

### Rare variant burden analysis

Single variant association testing is underpowered when variants are rare and a collapsing approach which aggregates variants by gene can be adopted to boost power. We extracted coding SNVs and indels with MAF < 0.01% in gnomAD^30^ annotated with one of the following: missense, in-frame insertion, in-frame deletion, start loss, stop gain, frameshift, splice donor, splice acceptor for each gene and further filtered them by CADD^23^ (v1.5) score using a threshold of ≥20 corresponding to the top 1% of all predicted deleterious variants in the genome. Variants meeting the following quality control filters were retained: MAC ≤ 20, median site-wide sequencing depth in non-missing samples > 20 and median GQ ≥ 30. Sample-level QC metrics for each site were set to minimum depth per sample of 10, minimum GQ per sample of 20 and ABratio P value > 0.001. Variants with significantly different missingness between cases and controls (P<10-5) or >5% missingness overall were excluded.

We employed the Scalable and Accurate Implementation of Generalized mixed model (SAIGE-GENE) (v0.42.1)^31^ to ascertain whether rare coding variation was enriched in cases on a per-gene basis exome-wide. SAIGE-GENE uses a generalized mixed-model to correct for population stratification and cryptic relatedness as well as a saddle point approximation and efficient resampling adjustment to account for the inflated type 1 error rates seen with unbalanced case-control ratios. It combines single-variant score statistics and their covariance estimate to perform SKAT-O^32^ gene-based association testing, upweighting rarer variants using the beta (1,25) weights option. SKAT-O is a combination of a traditional burden and variance-component test and provides robust power when the underlying genetic architecture is unknown. Sex and the top ten principal components were included as fixed effects when fitting the null model. A Bonferroni adjusted P value of 2.58×10^−6^ (0.05/19,364 genes) was used to determine the exome-wide significance threshold. Binary odds ratios and 95% confidence intervals were calculated for exome-wide significance genes by extracting the number of cases and controls carrying qualifying variants per gene in the collapsing analysis and applying a Fisher’s test in R.

### Validation of rare variant burden results in the UK Biobank

The AstraZeneca PheWAS portal (https://azphewas.com/) is a repository of gene-based phenotype associations derived from the UK Biobank, a prospective study over 500,000 individuals aged between 40-69 linking the health records with whole exome sequencing^33^. Collapsing gene-based analyses similar to our methodology were applied across the phenotypes for each gene. Twelve different sets of qualifying variant filters (models: ten dominant models, one recessive model, and one synonymous “control” variant model) were applied to test the association between 18,762 genes and 18,780 phenotypes after extensive quality control filters^34^. We queried the PheWas portal for gene associations in USD across all models (tag: “Source of report of N20 (calculus of kidney and ureter)”). Results were given by Fisher’s exact two-sided test P-values across each collapsed variant gene by model. The authors used a study-wide significance threshold of p ≤ 2×10^−09^.

### Polygenic risk scoring

In those cases, without a clear genetic diagnosis from the 100KGP clinical pipeline or a statistically significant gene association from the rare variant burden analysis, a polygenic risk score (PRS) was applied from a pre-validated, multi-ancestry PRS of USD ^35^. The PRS was applied to 336 cases and 24541 controls. The PRS included 7,670,833 markers derived from the UK Biobank using LDpred^36^. This was lifted over using the UCSC Liftover tool^37^ and imported into the 100KGP where the scoring was performed using the “score” command in PLINK2^38^. To test the significance between the PRS of cases, controls, and those with *SLC34A3* qualifying variants we applied a Kruskal-Wallis test followed by a paired Wilcox test to differentiate the source of statistical significance using base R functions. All plotting was performed with ggplot2 in R^39^. Odds ratios across PRS centiles were calculated with a logistic model using the generalised linear model tool in R; comparing phenotype with the centile of PRS using sex and the first ten principal components as covariates. Independence of PRS from the monoallelic *SLC34A3* signal was ascertained with a logistic model using phenotype and PRS with PRS*SLC34A3, presence or absence of a qualifying *SLC34A3* variant, sex and the first ten principal components as covariates. A null model containing just our covariates against the phenotype (sex and the first ten principal components) was used to calculate the model contribution to the phenotype variance. A liability adjusted Nagelkerke pseudoR^2^ was calculated for the model with and without the presence or absence of *SLC34A3* monoallelic variants using an estimated prevalence of USD of 5%^40,41^. Analysis of the generalized linear model outputs was performed with the pscl packages in R^42^.

## Results

### Participants

We identified 374 unrelated probands with USD (244 recruited to 100KGP under “nephrocalcinosis/nephrolithiasis” as their primary diagnosis and an additional 130 participants with HES codes indicating USD) and 24930 controls recruited to the UK 100,000 Genomes Project (100KGP).

Of the 244 primary recruited cases 26 were previously solved by 100KGP, with the relevant genetic diagnoses being reported back to the participants, representing a diagnostic yield of 10.7% (see table 1 for full breakdown). 21/244 (8.6%) had a primary diagnosis in keeping with stone forming disease whilst 5/244(2%) had other secondary diagnoses that were delivered to them via the clinical reporting pipeline that did not account for their stone disease. All disease-causing genes followed their established modes of inheritance.

**Table 1.** The 26 cases from the USD cohort who received a molecular diagnosis via the Genomics England Clinical interpretation portal. 21 cases were given a molecular diagnosis in keeping with USD whilst 5 patients were given molecular diagnoses that did not account for their urinary stone disease.

| Primary Diagnosis | Gene | No. of probands and zygosity |
| --- | --- | --- |
| Cystinuria | *SLC7A9* | 2 biallelic, 4 monoallelic |
|  | *SLC3A1* | 2 biallelic, 3 monoallelic |
| Primary Hyperoxaluria type 1 | *AGXT* | 3 biallelic |
| Primary Hyperoxaluria type 2 | *GRHPR* | 2 biallelic |
| Infantile hypercalcaemia | *CYP24A1* | 2 biallelic |
| Rain syndrome | *FAM20C* | 1 biallelic |
| Hereditary Hypophosphataemic Rickets with Hypercalciuria | *SLC34A3* | 1 biallelic |
| SLC34A1 associated USD | *SLC34A1* | 1 biallelic |
| Secondary diagnoses | Gene | No. of patients and zygosity |
| CHARGE syndrome | *CHD2* | 1 monoallelic |
| Alport syndrome | *COL4A4* | 1 monoallelic |
| Mucopolysaccharidosis | *GALNS* | 1 biallelic |
| Gitelman syndrome | *SLC12A3* | 1 biallelic |
| Beta Thalassemia | *HBB* | 1 monoallelic |

### Rare variant burden analysis of stone disease reveals significant enrichment in *SLC34A3*

Two genes showed statistically significant enrichment of rare variation in USD cases compared with controls: *SLC34A3* (P=*2*.*61×10*^*-07*^, OR=3.75, 95% CI 2.27-5.91) and *OR9K2*, encoding an olfactory receptor (P=*2*.*03×10*^*-06*^, OR = 8.47, 95 % CI 3.23-18.81) (figure 1). Full results of the association analysis can be found in Table S2.

**Figure 1.**
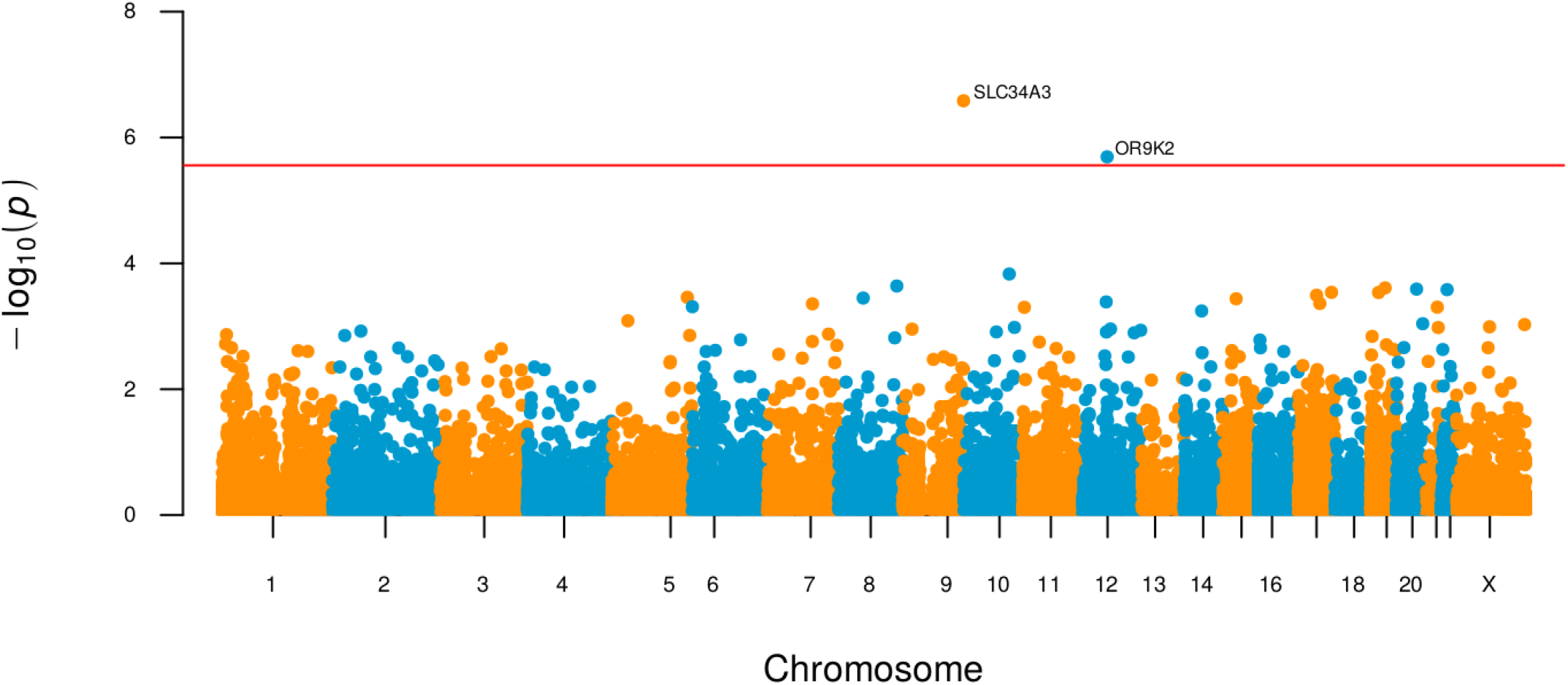
Gene based Manhattan plot of the SAIGE-GENE analysis with the missense+ tag. Each point is a gene made up of variants that are predicted to be at least as damaging as missense, with a CADD score >20 and have a minor allele frequency (MAF) <0.01% in the gnomAD database. The horizontal line indicates the threshold for exome-wide significance. The only exome-wide significant associations were with *SLC34A3* (P=2.61×10^−07^) and *OR9K2* (P= 2.03×10^−06^)

### Replication of results in the UK Biobank

Association of USD with rare variation in *SLC34A3*, but not *OR9K2*, was replicated in publicly available analyses of whole exome sequencing data from 3,147 cases and 255,496 controls within the UK Biobank, an independent dataset (https://azphewas.com/)^34^. In this analysis, multiple rare variants collapsing models were applied on a per gene basis and analysed with SKAT-O across all listed UK Biobank phenotypes. Under the “flexnonsynmtr” model, which equates to non-synonymous variants with a MAF<0.01% in both gnomAD and the UK Biobank with missense variants also having to fall within a region constrained for missense variation, USD was most strongly associated with *SLC34A3* (p*=3*.*67×10*^*-10*^, OR = 2.01, see figure 2).

**Figure 2.**
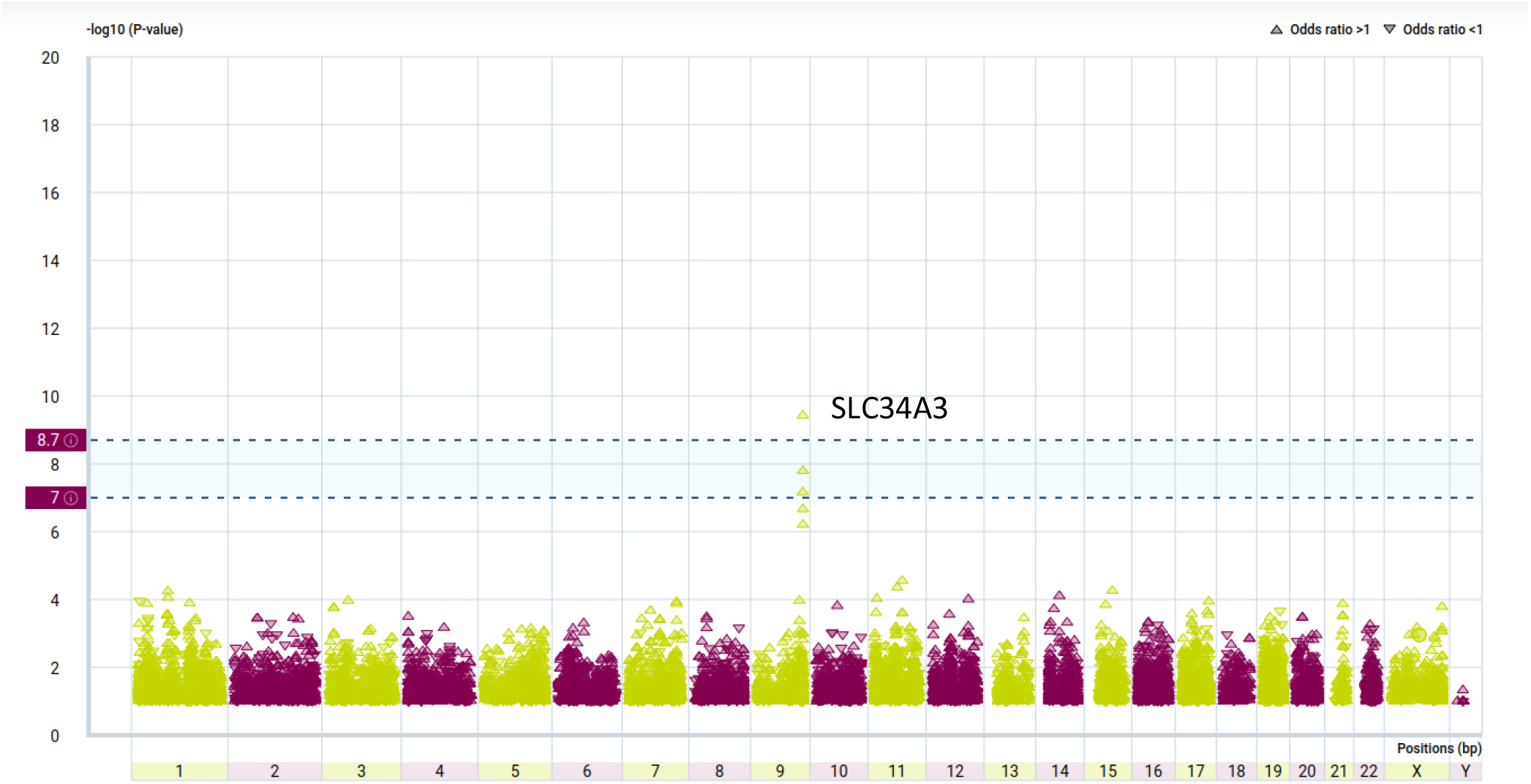
Gene based Manhattan plot of the UK Biobank analysis obtained from the AstraZeneca PheWas portal. Each symbol represents variants in a gene that are predicted to be non-synonymous with a minor allele frequency (MAF) <0.1% in both gnomAD and the UK Biobank. The only exome wide significant association was *SLC34A3* (p=3.67×10-10, OR = 2.01). The multiple symbols under SLC34A3 (the “build-up”) represent different analyses with respect to the predicted severity of the included variants. The lower dashed vertical line indicates exome-wide significance and the upper dashed line exome-wide significance corrected for the ∼1500 different phenotypes analysed.

### Phenotype/Genotype analysis of SLC43A3 cases

21 patients had qualifying variants that contributed to the *SLC34A3* association. 14/21 cases were recruited with stone disease, 1 with Congenital Anomalies of the Kidney or Urinary Tract, 4 with cystic kidney disease and two with intellectual disability. 19 patients were heterozygous, and 2 patients were compound heterozygous for qualifying *SLC34A3* variants with both of these cases’ variants being confirmed in *trans* (table 2, a full breakdown of the cases can be found in table S3). Qualifying variants were found in 6% of the ascertained stone population in the study (21/374) compared to 1.6% (389/24930) in the controls (all monoallelic).

**Table 2.** qualifying variants making up the *SLC34A3* association in the rare variant burden analysis using SAIGE-GENE.

| Variant type | Case Count (percentage) | Control Count (percentage) |
| --- | --- | --- |
| Missense | 17(75%) | 346(89%) |
| Frameshift | 1(4%) | 14(4%) |
| Splice Donor variant | 1(4%) | 20(5%) |
| Start lost | 1(4%) | 5(1.1%) |
| Stop gained | 3(13%) | 4(0.9%) |

**Table 3.**
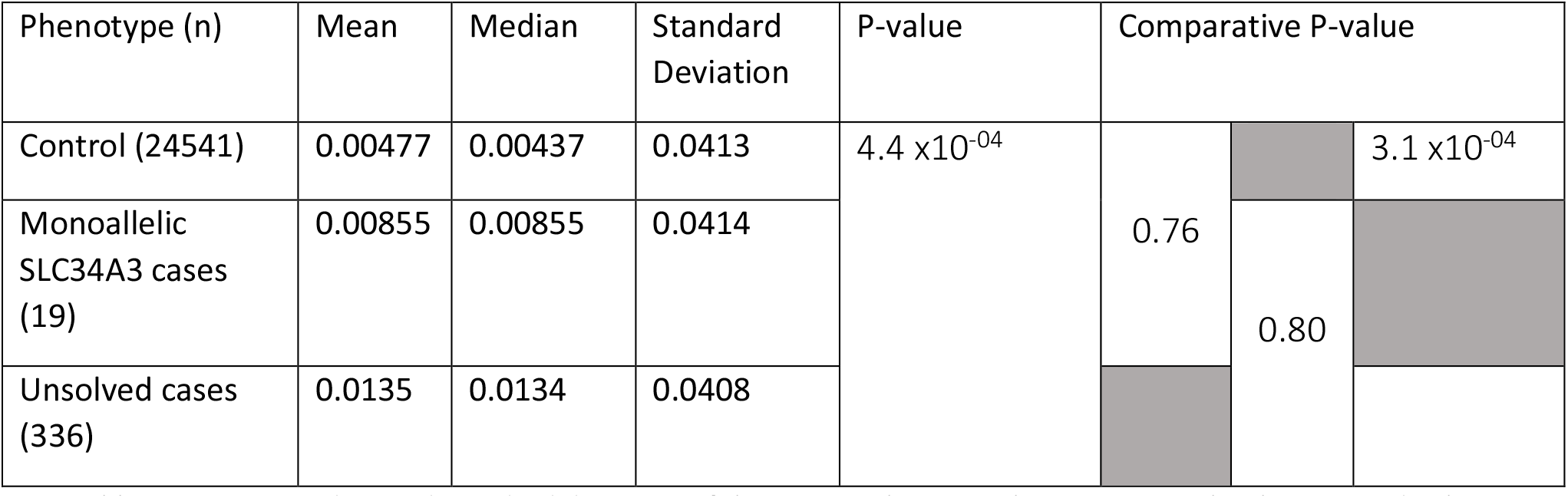
Mean, median and standard deviation of the stone polygenic risk score across the three tested cohorts. There were significant differences between the three cohorts by Kruskal-Wallis testing (p=4.4 x10-04). On further analysis with paired Wilcox testing this was found to be driven by the difference between controls and unsolved cases (p=3.1 x10-04). There was a near doubling of the *SLC34A3* cases mean and median compared to controls, but the numbers were too low to reach statistical significance.

All 21 patients had gone through the Genomics England clinical pipeline with 4 cases receiving genetic diagnoses including: *PKD2*-associated cystic kidney disease (two patients), Kabuki syndrome (*KMT2D*) and biallelic *SLC34A3*-associated USD. In the solved biallelic *SLC34A3* case both variants were annotated as (likely) pathogenic by Clinvar (rs199690076 and rs762710288) whereas in the unsolved biallelic *SLC34A3* case the evidence for a clinical grade diagnosis was weaker (rs369400414 and rs560440785). Whilst both variants met the inclusion criteria for collapsing rare variant association, they did not meet the benchmark for clinically reportable results. There is not enough phenotype data available for either biallelic *SLC34A3* case to ascertain whether they have the full gamut of Hereditary Hypophosphataemic Rickets with Hypercalciuria, but they do both have the hypercalciuria HPO code on record (full breakdown of HPO codes associated with each case in table S3).

The top ten HPO codes associated with the patients with *SLC34A3* associated USD are found in figure 3 below:

**Figure 3.**
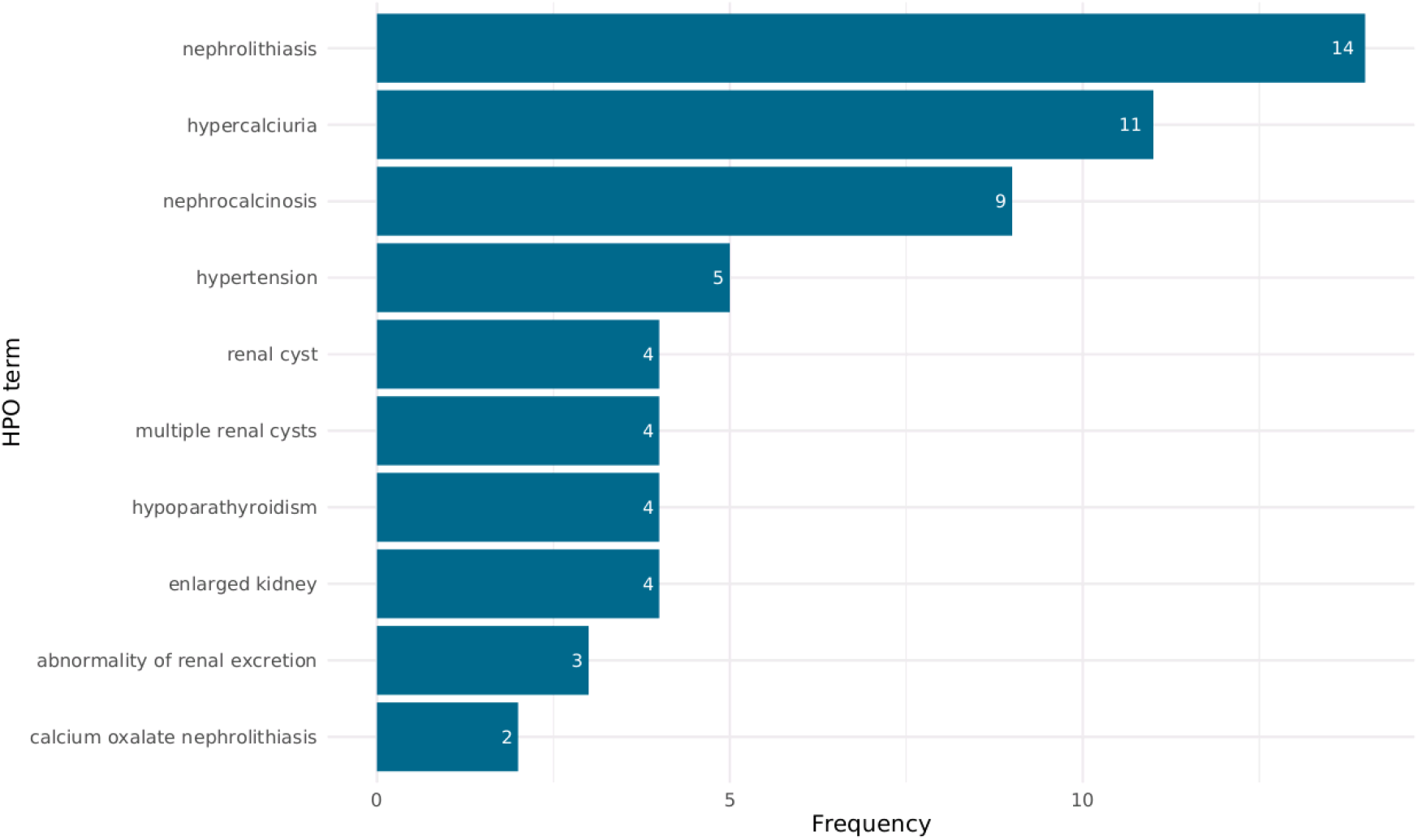
the top ten HPO codes associated with cases with qualifying *SLC34A3* variants. Nephrolithiasis makes up the most common associated clinical code followed by hypercalciuria and nephrocalcinosis.

### Polygenic risk scoring reveals significant difference between unsolved cases and controls

In cohorts of 336 unsolved cases and 24541 controls (both depleted for *SLC34A3* variants that would have qualified for inclusion in the SAIGE-GENE analysis), there was a significant elevation in USD PRS in unsolved cases compared with controls (P=3.1 *x10*^*-04*^). Initial analysis including a cohort of the cases with qualifying *SLC34A3* variants did identify statistically significant differences between the three cohorts (P=4.4 *x10*^*-04*^), but this signal was driven by the difference between unsolved cases and controls (figure 3). While the difference between the control group and the *SLC34A3* cases was large, it did not reach statistical significance given the small number of *SLC34A3* cases. Across the cohort, the PRS attributable risk of stone disease between cases and controls accounted for an odds ratio of 1.28. The upper bound 95% confidence interval for odds ratio of kidney stone disease among cases in the tenth decile of PRS was 2.31 (95% CI 1.12-3.51) (figure 4).

**Figure 4.**
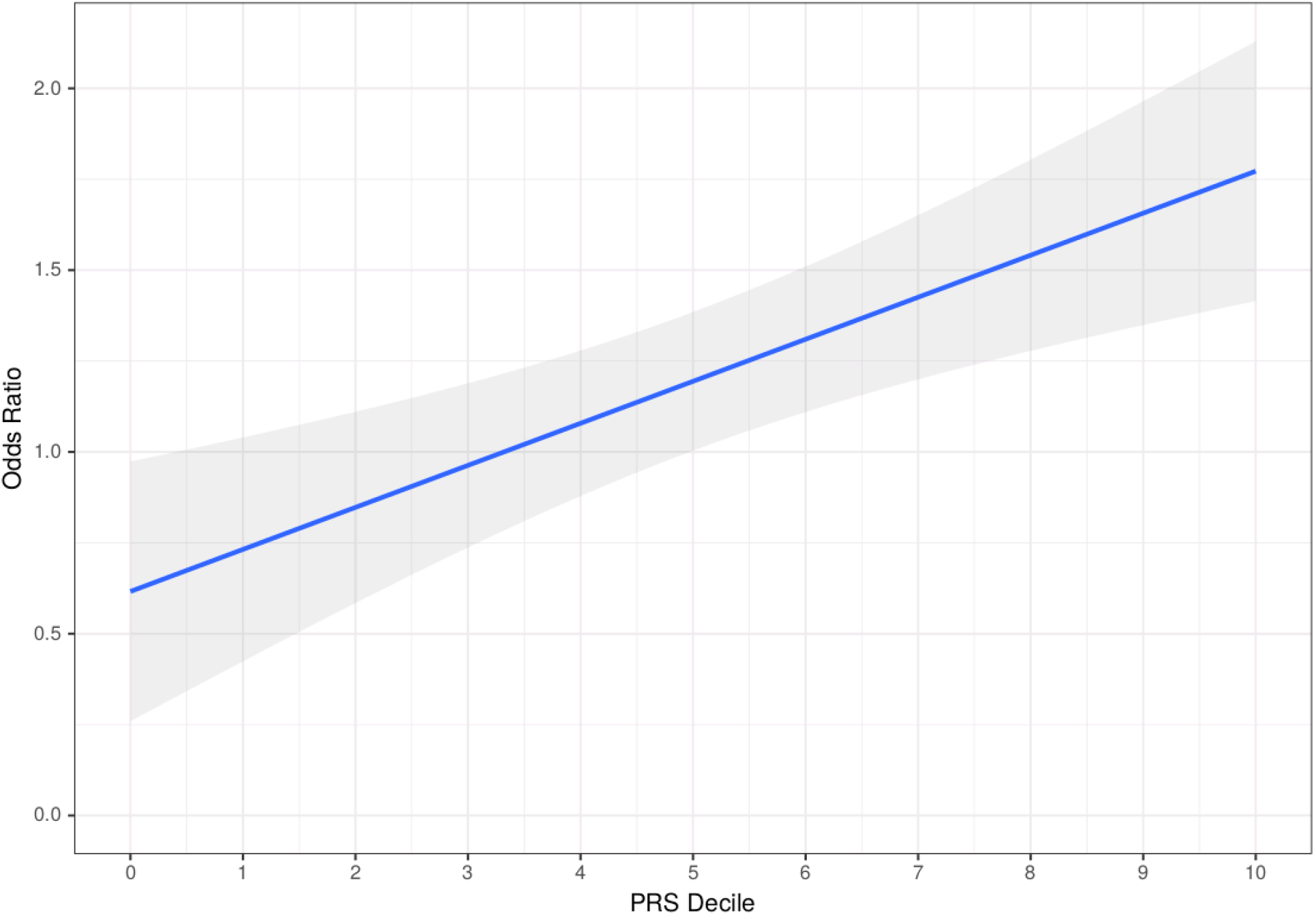
Linear regression plot of polygenic risk decile with the odds ratio of having urinary stone disease. The upper bound 95% confidence interval for the odds ratio of the top decile was associated with a 2.31 increased risk of urinary stone disease.

### The relationship between polygenic risk and monoallelic *SLC34A3* variants is independent and likely additive

In our model, there was a significant association between phenotype and both PRS (P=3.8×10^−04^) and the presence of a monoallelic *SLC34A3* variant (P=2.72×10^−08^). However, there was no significant association between the combined PRS/*SLC34A3* variable and the phenotype (P=0.77) implying independence between the *SLC34A3* and PRS signals.

The addition of the *SLC34A3* variant binary to the linear model including PRS led to a significant rise in the estimated variance explained by the model (liability adjusted pseudo-R^2^ rising from 5.1% to 14.2%) implying an additive effect of the *SLC34A3* monoallelic variants with the PRS and suggesting that rare monoallelic *SLC34A3* variants account for some of the gap between observed heritability of USD and that accounted for by common variants identified in previous GWAS.

## Discussion

We identified rare variants in *SLC34A3* and *OR9K2* as significantly associated with USD. *SLC34A3* encodes the sodium-dependent phosphate transport protein 2C expressed in the proximal tubule (NaPi-IIc). Data on the chemical composition of the stones in participants in the 100KGP are not available so it is possible that different (perhaps stronger) associations might be observable in sub-groups defined by stone composition. While the association with *SLC34A3* was independently replicated in the UK Biobank, a signal was not observed in this dataset at *OR9K2*, so the possibility exists that this finding is a type 1 error, which is well recognised with olfactory receptor genes owing to their enrichment for loss of function variation without clinical consequences^30,43^. However, similar olfactory gene associations have not been observed in other studies using the 100,000 Genomes Project dataset analyzed with similar methodology (ref 27 and unpublished observations), and there is increasing recognition that olfactory receptors regulate transport processes in many organ systems^44^: *OR9K2* is expressed in the intestine (Human Protein Atlas), and it is conceivable that it may be involved in the regulation of absorption of substrates with relevance to stone formation, such as oxalate or calcium. Therefore, further studies are needed to assess the relevance of *OR9K2* in USD.

Our results highlight the importance of *SLC34A3* as a contributor to USD, with more than 5% of patients in this cohort from the 100KGP harboring predicted damaging variants and independent replication of this association in the UK Biobank dataset^34^. Importantly, the odds ratio for stone disease with rare, predicted damaging variants in *SLC34A3* was higher than that conveyed by the top decile of a polygenic risk score derived from numerous common variants across the whole genome and a model combining PRS and *SLC34A3* monoallelic variants accounted for 14% of the genetic heritability of stone disease. This suggests that rare monoallelic variants in *SLC34A3* fall into an intermediate category of pathogenicity: they are insufficient to cause fully penetrant Mendelian disease but convey a higher disease risk than the aggregate effects of known common risk alleles. There is increasing recognition of such intermediate role for rare predicted damaging variants. For instance, approximately 1% of the general population carry such variants in *COL4A3* or *COL4A4* but they are not fully penetrant for the development of progressive chronic kidney disease (autosomal dominant Alport syndrome) and are therefore considered a risk factor ^45^.

In the AstraZeneca UK Biobank rare variant collapsing analysis that used twelve different sets of qualifying variant filters (models: ten dominant models, one recessive model, and one synonymous “control” variant model) *SLC34A3* was the gene significantly associated with USD with the lowest p-value. The association between *SLC34A3* and USD is strongest with models that include damaging missense variation and weakened if the filter is constrained to only those variants that are predicted to cause protein truncation, suggesting that even hypomorphic variants in this gene can contribute to the risk of USD.

Our findings therefore highlight the importance of monoallelic variants in *SLC34A3* for urinary calcium excretion and USD. *SLC34A3* was reported in 2006 as a recessive disease gene for the rare disorder hereditary hypophosphataemic rickets with hypercalciuria (HHRH)^46,47^. While there was already recognition in the original publication that heterozygous carriers in the affected families were frequently affected by hypercalciuria, it remains listed as a recessive disease gene in OMIM (*609826). Yet, there is good evidence for the impact of monoallelic variants: an investigation in a cohort of affected families showed that the risk of USD was 46%, 16% and 6% in subjects with biallelic, monoallelic or no causative variants, respectively^48^. This is consistent with a paradigm in which identification of a rare, deleterious monoallelic *SLC34A3* variants can be regarded as a risk factor for stone disease but is not a diagnostic finding.

The underlying mechanism is thought to be hypophosphataemia-mediated suppression of fibroblast growth factor-23(FGF23) with consequent activation of the 1-a hydroxylase and increased 1,25 dihydroxy vitamin D levels, which in turn stimulates intestinal calcium absorption^47^. The same mechanism is thought to apply in infantile hypercalcaemia due to biallelic loss-of-function variants in *SLC34A1*^49^. While the role of monoallelic *SLC34A1* variants in hypercalciuria has been controversial, large genome-wide studies have demonstrated a significant association between both coding and non-coding variants of *SLC34A1* and nephrolithiasis, consistent with the concept that a reduction in proximal tubular phosphate transport does increase the risk for kidney stones ^50,51,13^.

Our study provides evidence of clinical relevance for coding variants in *SLC34A3*, with a significant enrichment of rare and predicted deleterious variants in USD patients compared with controls, among participants of both the 100KGP and UK BioBank. While the 100KGP did not specifically encourage enrollment of patients with a family history of the respective disorders, it is possible that there may have been a recruitment bias that would have inflated the percentage of *SLC34A3*-related disease.

Nevertheless, the additional identification of rare *SLC34A3* variation as the strongest rare variant association in UK Biobank participants provides independent replication and raises the question of whether identification of these risk variants in individual patients would provide utility in clinical practice. While the modest risk affect precludes predictive use of such a test, the above pathophysiological mechanism suggests that phosphate supplementation may be a suitable treatment to stimulate FGF23 and thereby suppress 1-a hydroxylation of vitamin D in patients at risk of *SLC34A3*-related kidney stone disease. Indeed, successful use of this treatment has been reported^49,52,53^. However, clinical trial data would be needed to support such an intervention because there is a risk that large doses of phosphate supplementation increase the urinary phosphate concentrations with consequent increased risk of calcium phosphate precipitation. Indeed, nephrocalcinosis has been associated with phosphate supplementation in patients with *PHEX*-associated hypophosphataemic rickets, although these patients typically received enormous doses^54^. Thus, more data are needed before embarking on routine phosphate supplementation in *SLC34A3*-associated disease.

## Conclusion

Our study highlights the substantial contribution of rare and predicted damaging variants in *SLC34A3* to the burden of USD, helping to close the missing heritability gap and supporting the idea of routine genetic testing in affected patients.

## Data Availability

Genomic and phenotype data from participants recruited to the 100KGP can be accessed by application to Genomics England Ltd (https://www.genomicsengland.co.uk/about-gecip/joining-research-community/).
Code for the case-control ancestry-matching algorithm can be found at https://github.com/APLevine/PCA_Matching.

https://github.com/APLevine/PCA_Matching

## Data and code availability

Genomic and phenotype data from participants recruited to the 100KGP can be accessed by application to Genomics England Ltd (https://www.genomicsengland.co.uk/about-gecip/joining-research-community/).

Code for the case-control ancestry-matching algorithm can be found at https://github.com/APLevine/PCA_Matching.

## Data Availability

Genomic and phenotype data from participants recruited to the 100,000 Genomes Project can be accessed by application to Genomics England Ltd (https://www.genomicsengland.co.uk/about-gecip/joining-research-community/).

## Declaration of interests

The authors declare no competing interests.

## Supplemental Data

**Supplementary Appendix:** Genomics England Research Consortium

**Supplementary table 1** – list of terms used to create HES derived cohorts; list of codes used to exclude cases from controls

**Supplementary table 2** – full summary statistics from the rare variant burden analysis of stone cases versus controls using SAIGE-GENE

**Supplementary table 3** – full breakdown of the 21 cases that make up the SCL34A3 association using SAIGE-GENE.

**Supplementary figure 1 – Study Workflow.** The flowchart shows the number of samples included at each stage of filtering and the analytical strategies employed. MAF, minor allele frequency; Missense+, variants as least as damaging as a missense as per Ensembl’s the variant effect predictor tool; CADD, combined annotation dependent depletion score.

**Supplementary figure 2 – Ancestry matching.** Principal component analysis showing the first eight principal components for matched cases (red) and controls (green) and unmatched controls (grey).

**Supplementary figure 3 – PRS distributions.** Violin and boxplot showing the polygenic risk score distributions between controls, cases with qualifying SLC34A3 variants and unsolved patients.

## Acknowledgements

This research was made possible through access to the data and findings generated by the 100,000 Genomes Project. The 100,000 Genomes Project is managed by Genomics England Limited (a wholly owned company of the Department of Health and Social Care). The 100,000 Genomes Project is funded by the National Institute for Health Research and NHS England. The Wellcome Trust, Cancer Research UK and the Medical Research Council have also funded research infrastructure. The 100,000 Genomes Project uses data provided by patients and collected by the National Health Service as part of their care and support. The authors gratefully acknowledge the participation of the patients and their families recruited to the 100,000 Genomes Project.

OSA is funded by an MRC Clinical Research Training Fellowship (MR/S021329/1). MYC is funded by a Kidney Research UK Clinical Research Fellowship (TF_004_20161125). DPG is supported by the St Peter’s Trust for Kidney, Bladder and Prostate Research.

## Notes

### Competing Interest Statement

The authors have declared no competing interest.

### Author Declarations

Ethical approval for the 100KGP was granted by the Research Ethics Committee for East of England Cambridge South (REC Ref14/EE/1112).

