## Supplementary Data for "Rare variants in SLC34A3 explain missing heritability of urinary stone disease"

Variants in *SLC34A3* are a major contributor to urolithiasis – Supplementary appendix and figures

John C. Ambrose[1]; Prabhu Arumugam [1]; Roel Bevers [1]; Marta Bleda [1]; Freya Boardman-Pretty [1],2; Christopher R. Boustred [1]; Helen Brittain [1]; Mark J. Caulfield [1,2]; Georgia C. Chan [1]; Greg Elgar [1, 2]; Tom Fowler [1]; Adam Giess [1]; Angela Hamblin [1]; Shirley Henderson [1],2; Tim J. P. Hubbard [1]; Rob Jackson [1]; Louise J. Jones [1],2; Dalia Kasperaviciute [1, 2]; Melis Kayikci [1]; Athanasios Kousathanas [1]; Lea Lahnstein [1]; Sarah E. A. Leigh [1]; Ivonne U. S. Leong [1]; Javier F. Lopez [1]; Fiona Maleady-Crowe [1]; Meriel McEntagart [1]; Federico Minneci [1]; LoukasMoutsianas [1, 2]; Michael Mueller [1, 2]; Nirupa Murugaesu [1]; Anna C.Need [1, 2]; Peter O'Donovan [1]; Chris A.Odhams [1]; Christine Patch [1, 2]; Mariana Buongermino Pereira [1]; Daniel PerezGil [1]; John Pullinger [1]; Tahrima Rahim [1]; Augusto Rendon [1]; Tim Rogers [1]; Kevin Savage [1]; Kushmita Sawant [1]; Richard H. Scott [1]; Afshan Siddiq [1]; Alexander Sieghart [1]; Samuel C. Smith [1]; Alona Sosinsky [1, 2]; Alexander Stuckey [1]; Mélanie Tanguy [1]; Ana Lisa Taylor Tavares [1]; Ellen R. A. Thomas [1],2; Simon R. Thompson [1]; Arianna Tucci [1, 2]; Matthew J. Welland [1]; Eleanor Williams [1]; Katarzyna Witkowska [1, 2]; Suzanne M.Wood [1, 2].

1. Genomics England, London, UK

2. William Harvey Research Institute, Queen Mary University of London, London, EC1M 6BQ, UK.

### Supplementary figure 1

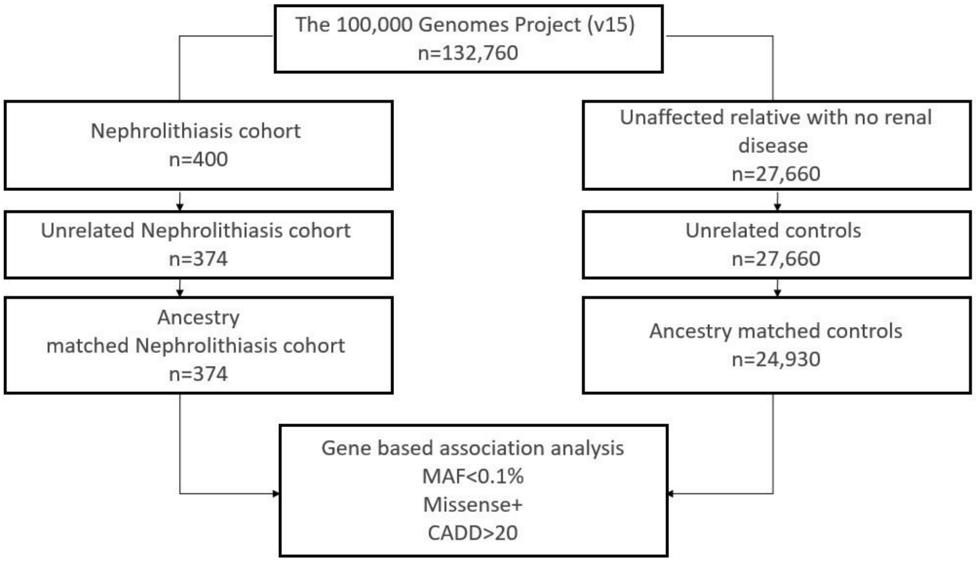


Supplementary figure 1 – Study Workflow. The flowchart shows the number of samples included at each stage of filtering and the analytical strategies employed. MAF, minor allele frequency; Missense+, variants as least as damaging as a missense as per Ensembl's the variant effect predictor tool; CADD, combined annotation dependent depletion score.

### Supplementary figure 2

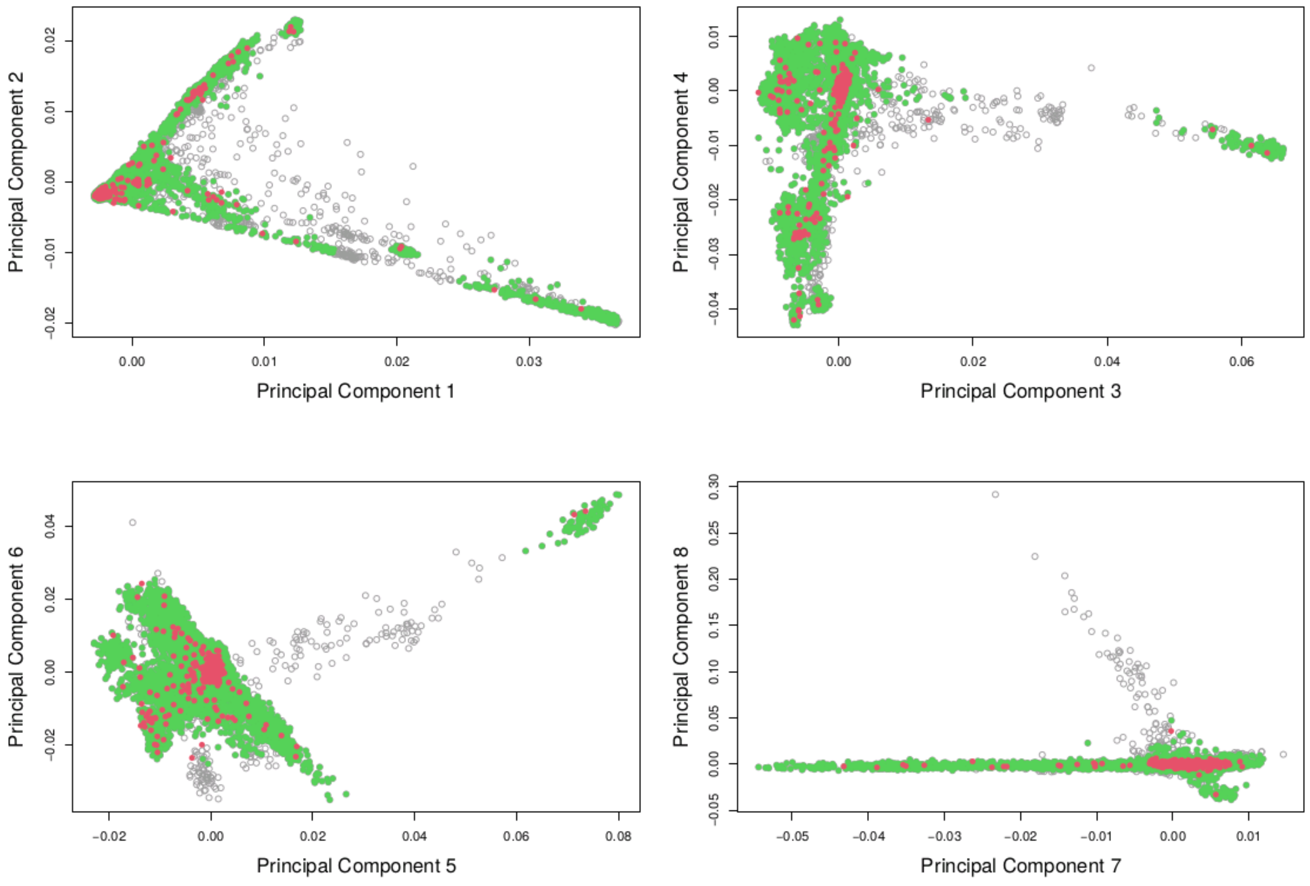


Supplementary figure 2 – Ancestry matching. Principal component analysis showing the first eight principal components for matched cases (red) and controls (green) and unmatched controls (grey). This highlights that cases are taken from multiple different ancestries with the appropriate matched controls.

### Supplementary figure 3

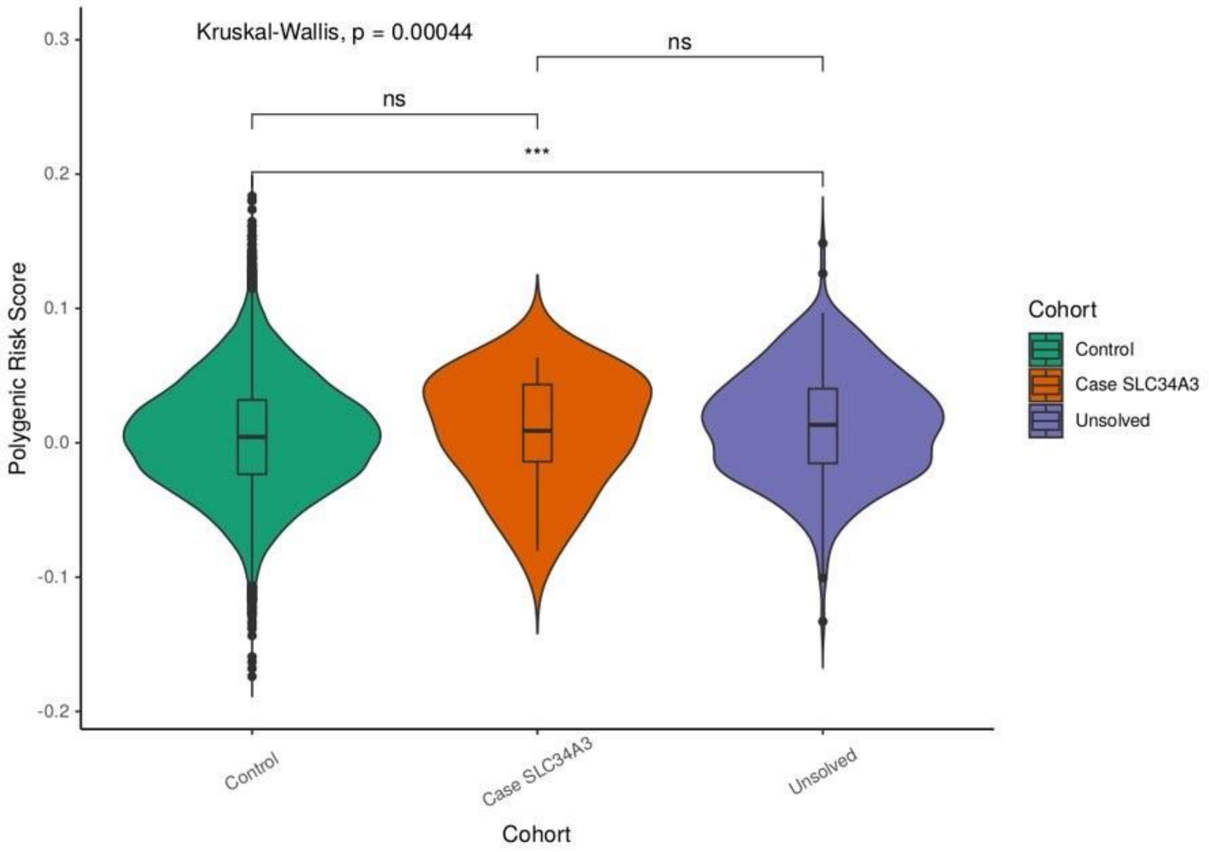


Supplementary figure 3 - Violin and boxplot showing the polygenic risk score (PRS) distributions between controls (those with qualifying SLC34A3 variants removed), cases with qualifying SLC34A3 variants and unsolved patients who have neither a reportable variant or a qualifying SLC34A3. The means of the three PRS were compared with a Kruskal-Wallis test (p=4.4 $x10^{-04}$) with the signal being driven by the difference between unsolved cases and controls (paired Willcox =3.1 $x10^{-04}$). **** = statistical significance, ns = no significant difference*
